# Potential association of HLA and KIR genetic profiles with resistance to HIV infection in high-risk men who have sex with men

**DOI:** 10.64898/2026.04.30.26352161

**Authors:** Ana C. Ossa-Giraldo, Yurany Blanquiceth, Lizdany Flórez-Álvarez, Adrián Peñata, Julián Bustamante, Nancy D. Marin, Winston Rojas, Juan C. Hernandez, Wildeman Zapata-Builes

## Abstract

Understanding the immune response against HIV-1 and the natural resistance exhibited by HIV-exposed Seronegative Individuals (HESN) offers the possibility of proposing new control strategies. Several studies suggest an important role of HLA and KIR genes in protecting against HIV-1 infection. Moreover, there is an important gap in the knowledge of these genetic factors in seronegative Latin American men who have sex with men (MSM), a population largely underrepresented in HIV immunogenetic studies.

This study aimed to identify HLA and KIR genetic profile associated with potential resistance to HIV-1 acquisition, in a cross-sectional study including a cohort of 60 HIV-1-seronegative Colombian MSM at low and high risk of HIV-1 infection.

The high-risk group showed a higher frequency of the HLA-B*18 allele, and a lower frequency of the HLA*B35, which have been previously associated with protection and susceptibility to HIV-1 infection respectively. Likewise, the high-risk group exhibited a low frequency of Bx haplotypes, a higher frequency of one AA haplotype and differences in KIR gene profile, with a low frequency of the inhibitory KIR2DL5 and both activating KIR2DS1, KIR2DS2 and KIR2DS5 genes. These findings suggest that host immunogenetic factors may contribute to resistance to HIV-1 acquisition in highly exposed individuals.

## Introduction

HIV-exposed seronegative individuals (HESN) are small populations who, although repeatedly exposed to the virus, do not present serological or clinical evidence of HIV infection^1^. Understanding the immune factors associated with natural resistance to HIV-1 in this population is significant in identifying the mechanisms determining the transmission and establishment of HIV infection^2^. However, the study of HIV-exposed seronegative (HESN) individuals is challenging due to their low frequency in the general population^2^ and the success of HIV prevention and treatment strategies, including pre-exposure prophylaxis (PrEP) and the “undetectable = untransmittable” (U=U) paradigm^3,4^. While these advances are highly beneficial for public health, they make it increasingly difficult to identify individuals who remain seronegative despite sustained, high-risk HIV exposure—a key criterion for HESN research. In this context, men who have sex with men (MSM) who engage in high-risk sexual behaviors (e.g., unprotected anal intercourse, multiple concurrent partners, and substance use-associated transactional sex) remain a relevant population for identifying potential HESN individuals, particularly in settings where HIV prevalence among MSM is high, such as Colombia (7.6% worldwide and approximately 15% in Colombia)^5^. Natural killer (NK) cells are key components of the innate immune response, capable of rapidly reacting to viral infections through the integrated signaling of activating and inhibitory surface receptors^6^. They play an important role during the early stages of HIV infection^6^. Although NK responses were initially considered non-specific, accumulating evidence indicates that these cells can develop memory-like properties against viral antigens, including HIV^7^. During acute infection, NK cells expand and exhibit functional responses against HIV, both in vivo and in vitro settings^8^. Additionally, NK cell-mediated antibody-dependent cellular cytotoxicity (ADCC) and interactions between specific human leukocyte antigen (HLA) alleles and killer immunoglobulin-like receptors (KIRs) have been linked to improved clinical outcomes^9^. In this sense, the immune response against HIV-1 can be influenced by genetic background. Specific HLA and KIR genes have been associated with natural resistance and susceptibility to HIV-1 infection and progression although they differ across populations^10,11^.

While some HLA class I (HLA-I) alleles have been associated with decreased risk of infection in HESN, and infection control in elite controllers, others have been associated with increased susceptibility to HIV-1 infection^12–14^. For instance, HLA-I A*01, A*24, A*25, A*32, B*13, B*14, B*18, B*44, B*27, B*57, B*58, C*06 and C*07 alleles have been associated with natural resistance to HIV-1^12,15–18^. In contrast, the HLA B*35 and B*07 alleles have been frequently associated with susceptibility and rapid progression to AIDS^19–21^.

KIR are highly polymorphic activating and inhibitory receptors that bind HLA-A, HLA-B, and HLA-C molecules to regulate NK cell functions. Specifically, KIR and HLA alleles and haplotypes influence NK cell responses against HIV-1 by activating and inhibitory signals on NK cells through their respective HLA ligands on infected cells^22,23^. Activating KIR, mainly KIR3DS1, and its ligand HLA-Bw4 are commonly associated with protection against HIV-1 in HESN cohorts^24,25^. Interestingly, inhibitory KIR genes such as KIR3DL1, and the KIR2DL2/KIR2DL3 heterozygosity expressed in the absence of HLA-C1 have also been associated with protection against HIV-1^26,27^. The strong association between KIR and HLA suggests that susceptibility or resistance during HIV-1 exposure could be mediated by the genetic background of each individual.

Most information about KIR genes and their protective role in HIV-1 was derived from studies with Caucasian, African, and Asian populations. However, there is a significant gap in the existing knowledge regarding the role of KIR-HLA genes in protecting against HIV-1 in Latin-American individuals^16,28^. This study aimed to determine the HLA and KIR genes, and KIR haplotypes associated with HIV-1 protection in a cohort of Colombian seronegative MSM at low and high risk of HIV-1 infection.

## Materials and Methods

### Study design and population

A cross-sectional study was conducted on 60 seronegative men who have sex with men (MSM) from Medellín, Colombia. These participants were selected from a cohort previously described^29,30^. All subjects were Hispanic/Latino. Individuals who were sex workers, using pre-exposure prophylaxis (PrEP), or homozygous for the delta 32 CCR5 mutation were excluded. All participants were negative for anti-HIV-1/2 antibodies and HIV-1 proviral DNA. Participants were classified as high-risk or low-risk for HIV acquisition based on their sexual behaviors. MSM in the high-risk group had a higher frequency of sexual partners in the last 3 months before the study participation (median 30 vs. 2), unprotected anal intercourse in the same period (median 12.5 vs. 2) and lifetime sexual partners (median 1,708 vs. 26). The study was conducted in accordance with the Declaration of Helsinki and approved by the Ethics Committee of the University of Antioquia School of Medicine (Act No. 007, May 22, 2014). Informed consent was obtained from all participants prior to their inclusion in the study.

### HLA and KIR genotyping

Peripheral blood was used to extract Genomic DNA, and the concentration was adjusted to 40ng/ul. HLA-A and B alleles were amplified using Rapid HLA-A, B, SSO typing kit (Immucor-Lifecodes System, Inc Stanford, CT). The hybridization of the PCR products was carried out in the liquid phase using Luminex® beads, which recognize the polymorphism of the second and third exon on HLA class I molecules. Luminex® IS 200 System, xPONENT® 3.1 Software (Luminex Corporation, Austin, TX) was used to read the assay, and HLA typing was analyzed with Match IT™ DNA software (Immucor-Lifecodes System, Inc Stanford, CT).

KIR genotyping was performed on 55 individuals (high-risk, n=14; low-risk, n=41) because DNA was not available for the remaining five participants from the original cohort of 60. KIR genotyping was performed by PCR-SSOP (Polymerase Chain Reaction-Sequence Specific Oligonucleotide Probes) using the SSO LabType® kit (One Lambda, San Diego, CA, USA) and following the manufacturer’s instructions. DNA integrity and purity was measured in NanoDrop (Thermo Scientific) and the concentration was adjusted to 20 ng/µl of genomic DNA per 20µl final volume of PCR reaction. The amplified product was hybridized to microspheres containing probes specific to the KIR gene. The resulting products were analyzed using the Luminex® IS 200 System flow cytometer (Luminex Corporation, Austin, TX) and final KIR typing was analyzed with the HLA Fusion™ Software (One Lambda, San Diego, CA, USA). The results were compared by matching the pattern of positive and negative beads for the specific genes with the information in the product worksheet. Fourteen KIR genes and two pseudogenes were assessed: KIR3DL3, KIR2DS2, KIR2DL3, KIR2DL2, KIR2DS3, KIR2DP1, KIR2DL1, KIR3DP1, KIR2DL4, KIR3DL1, KIR3DS1, KIR2DL5, KIR2DS5, KIR2DS1, KIR2DS4 and KIR3DL2.

### KIR haplotypes

Based on gene content and data from the Allele Frequency Net Database (http://allelefrequencies.net/default), KIR genotypes were classified into two categories: AA haplotypes, which carry only genes from the A haplotype, and Bx haplotypes, which include genotypes with genes from both A and B haplotypes (AB) or exclusively from the B haplotype (BB)^31,32^.

### Statistical Analysis

Differences between groups were assessed using Chi-square tests for categorical variables and, after normality testing, parametric or non-parametric tests for continuous variables. The strength of association between each genetic marker and the protection against HIV-1 acquisition was estimated using odds ratios (OR) with 95% confidence intervals (CI). Crude and adjusted ORs were calculated for each independent variable.

Given the exploratory nature of this study and the exceptional difficulty of recruiting potential HIV-exposed seronegative (HESN) individuals with extreme risk behavior, the sample size is inherently limited. Applying a stringent multiple-comparison correction (e.g., Bonferroni) would increase the risk of type II errors (false negatives), potentially masking biologically relevant associations that are consistent with prior HESN literature. Therefore, we report nominal p-values (p < 0.05) without correction, and all findings should be considered hypothesis-generating. The Linkage disequilibrium (LD) and haplotype analysis were performed using Haploview software (Copyright (c) 2003-2006 Broad Institute of MIT and Harvard (https://sourceforge.net/projects/haploview/). All other statistical analyses were conducted using SPSS, version 25.0 (IBM Corp. Released 2017. IBM SPSS Statistics for Windows, Version 25.0. Armonk, NY: IBM Corp).

## Results

### Sociodemographic and sexual behavior data of MSM

Sociodemographic characteristics and sexual behaviors of both MSM groups have been previously described^29,30^. Briefly, most participants in both groups self-identified as gay/homosexual males, were single, and had access to vocational or professional education. Notably, despite being seronegative for HIV-1 and lacking proviral DNA, the high-risk group exhibited sexual risk behaviors strongly associated with HIV-1 seroconversion in MSM cohorts^29^. Compared with the low-risk group, the high-risk group reported a significantly higher median number of lifetime sexual partners (1,078 vs. 26; p < 0.001), a higher median number of unprotected sexual intercourse in the preceding three months (10 vs. 2; p = 0.001), and a greater proportion of individuals who reported never using a condom with their regular partners (93.8% vs. 75.0%; p = 0.001). Furthermore, 81.3% of high-risk MSM reported never using a condom with their casual partners, and 87.5% reported a history of sexually transmitted infections. Thus, this cohort represents an exceptional population whose seronegativity defies their extreme risk profile.

### The high-risk MSM group exhibited a higher frequency of protective HLA alleles

HLA-A and HLA-B alleles were determined in both study groups. The high-risk group showed a significantly higher frequency of HLA-B*18 compared with the low-risk group (25.00% vs. 4.50%, p = 0.036; OR = 0.143, 95% CI 0.023-0.877) and a significantly lower frequency of HLA-B*35 (6.30% vs. 36.40%, p = 0.047; OR = 8.571, 95% CI 1.034-71.081). No statistically significant differences were observed for HLA-A alleles or for the HLA-Bw4 epitope (Table 1). The HLA-A and HLA-B genotypes are provided in Supplementary Data 1.

**Table 1.**
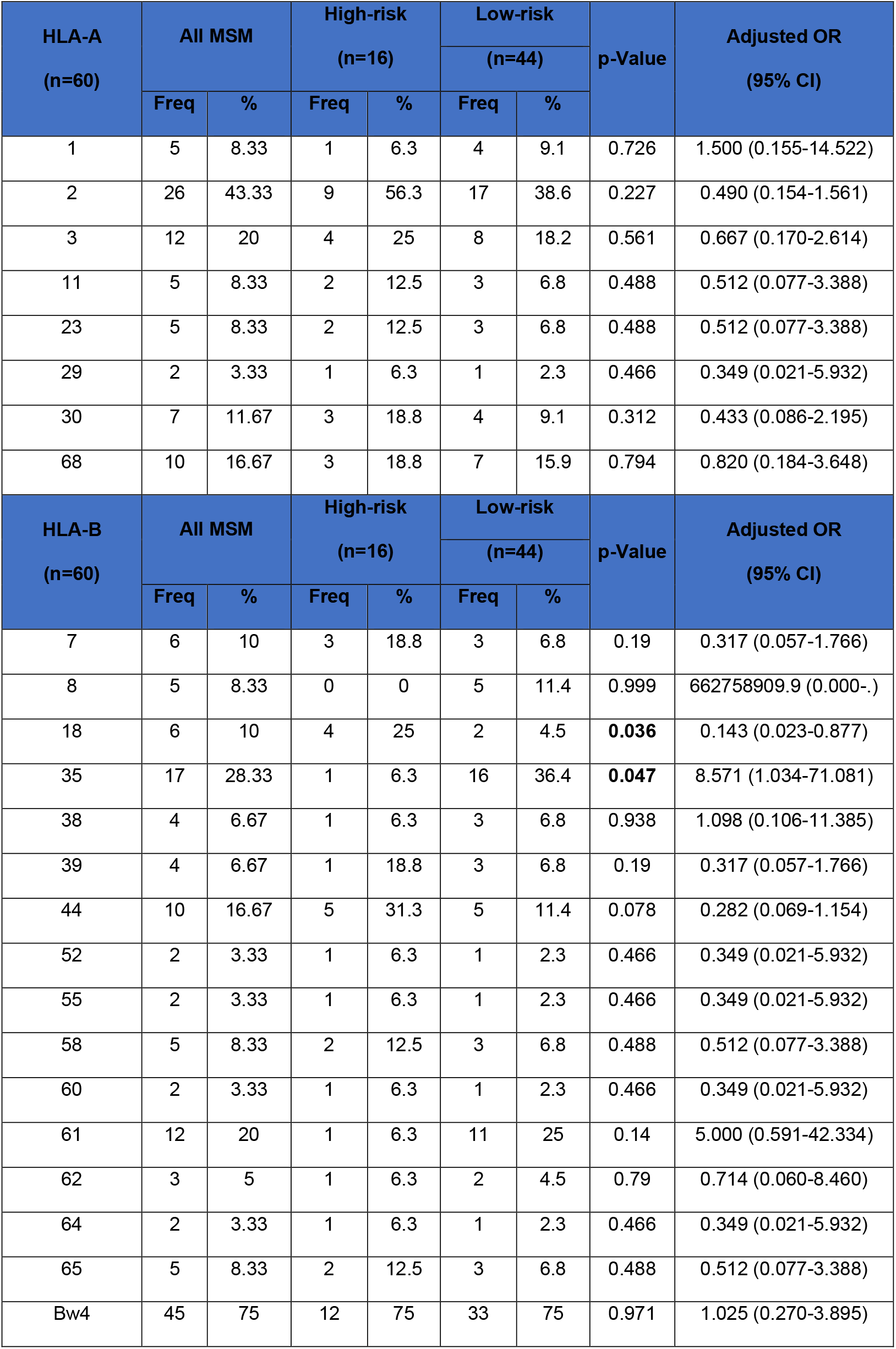
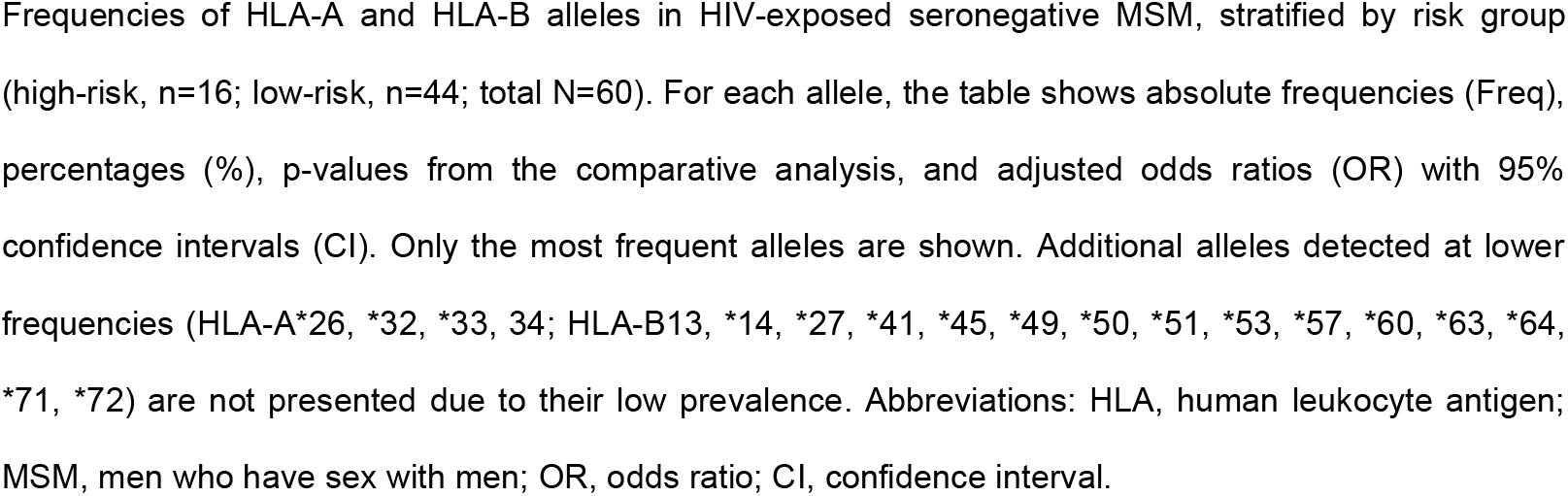
HLA alleles frequency in MSM groups.

| HLA-A<br>(n=60) | All MSM |  | High-risk<br>(n=16) |  | Low-risk |  | p-Value | Adjusted OR<br>(95% CI) |
| --- | --- | --- | --- | --- | --- | --- | --- | --- |
|  |  |  |  |  | (n=44) |  |  |  |
|  | Freq | % | Freq | % | Freq | % |  |  |
| 1 | 5 | 8.33 | 1 | 6.3 | 4 | 9.1 | 0.726 | 1.500 (0.155-14.522) |
| 2 | 26 | 43.33 | 9 | 56.3 | 17 | 38.6 | 0.227 | 0.490 (0.154-1.561) |
| 3 | 12 | 20 | 4 | 25 | 8 | 18.2 | 0.561 | 0.667 (0.170-2.614) |
| 11 | 5 | 8.33 | 2 | 12.5 | 3 | 6.8 | 0.488 | 0.512 (0.077-3.388) |
| 23 | 5 | 8.33 | 2 | 12.5 | 3 | 6.8 | 0.488 | 0.512 (0.077-3.388) |
| 29 | 2 | 3.33 | 1 | 6.3 | 1 | 2.3 | 0.466 | 0.349 (0.021-5.932) |
| 30 | 7 | 11.67 | 3 | 18.8 | 4 | 9.1 | 0.312 | 0.433 (0.086-2.195) |
| 68 | 10 | 16.67 | 3 | 18.8 | 7 | 15.9 | 0.794 | 0.820 (0.184-3.648) |
| HLA-B<br>(n=60) | All MSM |  | High-risk<br>(n=16) |  | Low-risk |  | p-Value | Adjusted OR<br>(95% CI) |
|  |  |  |  |  | (n=44) |  |  |  |
|  | Freq | % | Freq | % | Freq | % |  |  |
| 7 | 6 | 10 | 3 | 18.8 | 3 | 6.8 | 0.19 | 0.317 (0.057-1.766) |
| 8 | 5 | 8.33 | 0 | 0 | 5 | 11.4 | 0.999 | 662758909.9 (0.000-.) |
| 18 | 6 | 10 | 4 | 25 | 2 | 4.5 | **0.036** | 0.143 (0.023-0.877) |
| 35 | 17 | 28.33 | 1 | 6.3 | 16 | 36.4 | **0.047** | 8.571 (1.034-71.081) |
| 38 | 4 | 6.67 | 1 | 6.3 | 3 | 6.8 | 0.938 | 1.098 (0.106-11.385) |
| 39 | 4 | 6.67 | 1 | 18.8 | 3 | 6.8 | 0.19 | 0.317 (0.057-1.766) |
| 44 | 10 | 16.67 | 5 | 31.3 | 5 | 11.4 | 0.078 | 0.282 (0.069-1.154) |
| 52 | 2 | 3.33 | 1 | 6.3 | 1 | 2.3 | 0.466 | 0.349 (0.021-5.932) |
| 55 | 2 | 3.33 | 1 | 6.3 | 1 | 2.3 | 0.466 | 0.349 (0.021-5.932) |
| 58 | 5 | 8.33 | 2 | 12.5 | 3 | 6.8 | 0.488 | 0.512 (0.077-3.388) |
| 60 | 2 | 3.33 | 1 | 6.3 | 1 | 2.3 | 0.466 | 0.349 (0.021-5.932) |
| 61 | 12 | 20 | 1 | 6.3 | 11 | 25 | 0.14 | 5.000 (0.591-42.334) |
| 62 | 3 | 5 | 1 | 6.3 | 2 | 4.5 | 0.79 | 0.714 (0.060-8.460) |
| 64 | 2 | 3.33 | 1 | 6.3 | 1 | 2.3 | 0.466 | 0.349 (0.021-5.932) |
| 65 | 5 | 8.33 | 2 | 12.5 | 3 | 6.8 | 0.488 | 0.512 (0.077-3.388) |
| Bw4 | 45 | 75 | 12 | 75 | 33 | 75 | 0.971 | 1.025 (0.270-3.895) |

Frequencies of HLA-A and HLA-B alleles in HIV-exposed seronegative MSM, stratified by risk group (high-risk, n=16; low-risk, n=44; total N=60). For each allele, the table shows absolute frequencies (Freq), percentages (%), p-values from the comparative analysis, and adjusted odds ratios (OR) with 95% confidence intervals (CI). Only the most frequent alleles are shown. Additional alleles detected at lower frequencies (HLA-A\*26, \*32, \*33, 34; HLA-B13, \*14, \*27, \*41, \*45, \*49, \*50, \*51, \*53, \*57, \*60, \*63, \*64, \*71, \*72) are not presented due to their low prevalence. Abbreviations: HLA, human leukocyte antigen; MSM, men who have sex with men; OR, odds ratio; CI, confidence interval.

### MSM in the high-risk group exhibited protective KIR genes & haplotypes

The presence of 14 KIR genes and 2 pseudogenes was determined. Compared with the low-risk group, the high-risk group exhibited significantly lower frequencies of the activating KIR genes 2DS1 (7.14% vs. 48.78%, p = 0,030; OR = 15.000, 95% CI 1.759-127,913), 2DS2 (28.57% vs. 63.41%, p = 0.022; OR = 4.727, 95% IC 1.175-19.016) and 2DS5 (7.14% vs. 43.90%, p = 0.015; OR = 9.900, 95% CI, 1.160-84.471), as well as a lower frequency of the inhibitory KIR gene 2DL5 (7.14% vs. 46.34%, p = 0.011; OR = 17,273, 95% CI 1.942-153.664) (Table 2).

**Table 2.**
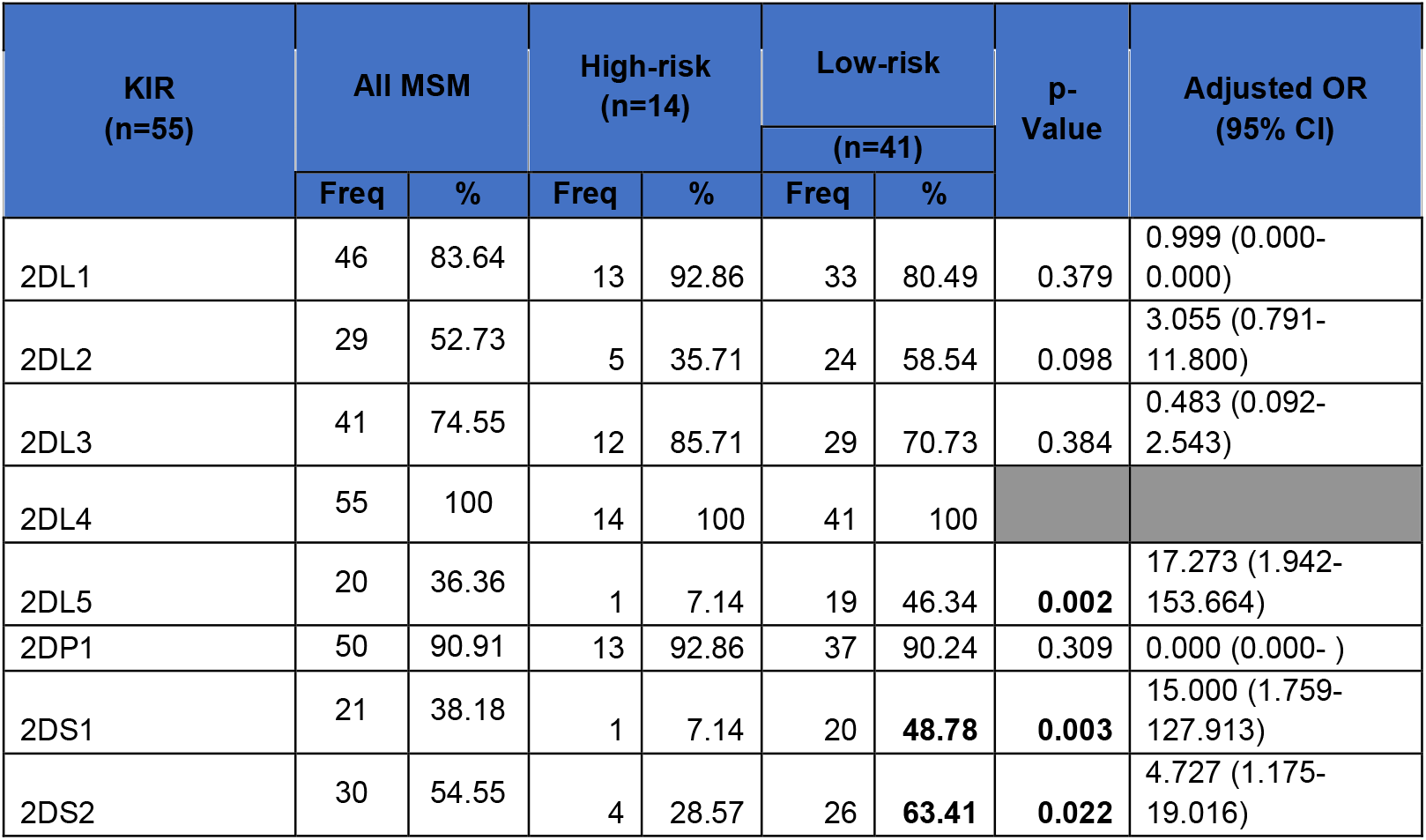
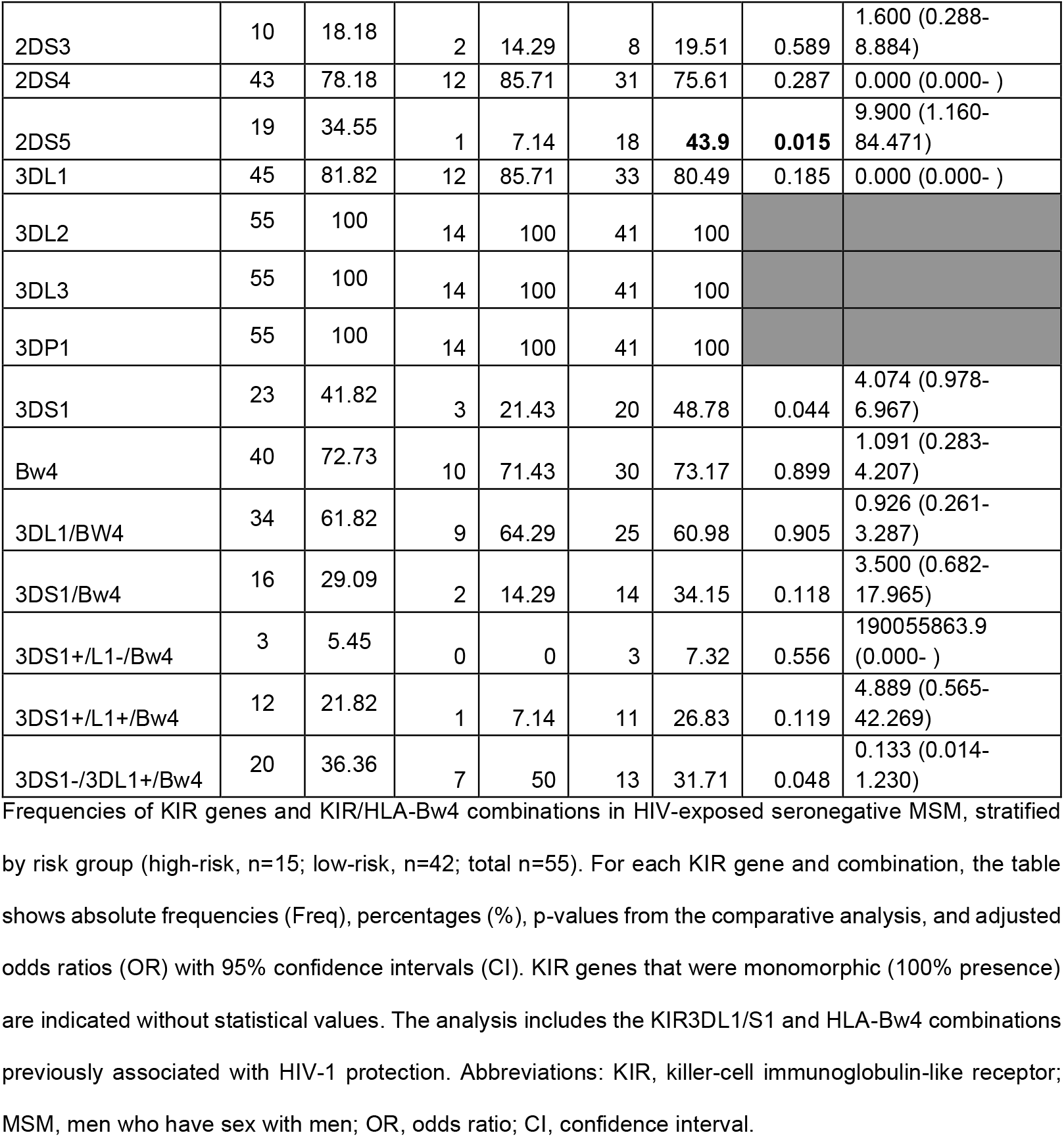
KIR genes frequency in MSM groups.

| KIR<br>(n=55) | All MSM |  | High-risk<br>(n=14) |  | Low-risk<br>(n=41) |  | p-Value | Adjusted OR<br>(95% CI) |
| --- | --- | --- | --- | --- | --- | --- | --- | --- |
|  | Freq | % | Freq | % | Freq | % |  |  |
| 2DL1 | 46 | 83.64 | 13 | 92.86 | 33 | 80.49 | 0.379 | 0.999 (0.000-0.000) |
| 2DL2 | 29 | 52.73 | 5 | 35.71 | 24 | 58.54 | 0.098 | 3.055 (0.791-11.800) |
| 2DL3 | 41 | 74.55 | 12 | 85.71 | 29 | 70.73 | 0.384 | 0.483 (0.092-2.543) |
| 2DL4 | 55 | 100 | 14 | 100 | 41 | 100 |  |  |
| 2DL5 | 20 | 36.36 | 1 | 7.14 | 19 | 46.34 | **0.002** | 17.273 (1.942-153.664) |
| 2DP1 | 50 | 90.91 | 13 | 92.86 | 37 | 90.24 | 0.309 | 0.000 (0.000- ) |
| 2DS1 | 21 | 38.18 | 1 | 7.14 | 20 | **48.78** | **0.003** | 15.000 (1.759-127.913) |
| 2DS2 | 30 | 54.55 | 4 | 28.57 | 26 | **63.41** | **0.022** | 4.727 (1.175-19.016) |
| 2DS3 | 10 | 18.18 | 2 | 14.29 | 8 | 19.51 | 0.589 | 1.600 (0.288-8.884) |
| 2DS4 | 43 | 78.18 | 12 | 85.71 | 31 | 75.61 | 0.287 | 0.000 (0.000- ) |
| 2DS5 | 19 | 34.55 | 1 | 7.14 | 18 | **43.9** | **0.015** | 9.900 (1.160-84.471) |
| 3DL1 | 45 | 81.82 | 12 | 85.71 | 33 | 80.49 | 0.185 | 0.000 (0.000- ) |
| 3DL2 | 55 | 100 | 14 | 100 | 41 | 100 |  |  |
| 3DL3 | 55 | 100 | 14 | 100 | 41 | 100 |  |  |
| 3DP1 | 55 | 100 | 14 | 100 | 41 | 100 |  |  |
| 3DS1 | 23 | 41.82 | 3 | 21.43 | 20 | 48.78 | 0.044 | 4.074 (0.978-6.967) |
| Bw4 | 40 | 72.73 | 10 | 71.43 | 30 | 73.17 | 0.899 | 1.091 (0.283-4.207) |
| 3DL1/Bw4 | 34 | 61.82 | 9 | 64.29 | 25 | 60.98 | 0.905 | 0.926 (0.261-3.287) |
| 3DS1/Bw4 | 16 | 29.09 | 2 | 14.29 | 14 | 34.15 | 0.118 | 3.500 (0.682-17.965) |
| 3DS1+/L1-/Bw4 | 3 | 5.45 | 0 | 0 | 3 | 7.32 | 0.556 | 190055863.9 (0.000- ) |
| 3DS1+/L1+/Bw4 | 12 | 21.82 | 1 | 7.14 | 11 | 26.83 | 0.119 | 4.889 (0.565-42.269) |
| 3DS1-/3DL1+/Bw4 | 20 | 36.36 | 7 | 50 | 13 | 31.71 | 0.048 | 0.133 (0.014-1.230) |

Frequencies of KIR genes and KIR/HLA-Bw4 combinations in HIV-exposed seronegative MSM, stratified by risk group (high-risk, n=15; low-risk, n=42; total n=55). For each KIR gene and combination, the table shows absolute frequencies (Freq), percentages (%), p-values from the comparative analysis, and adjusted odds ratios (OR) with 95% confidence intervals (CI). KIR genes that were monomorphic (100% presence) are indicated without statistical values. The analysis includes the KIR3DL1/S1 and HLA-Bw4 combinations previously associated with HIV-1 protection. Abbreviations: KIR, killer-cell immunoglobulin-like receptor; MSM, men who have sex with men; OR, odds ratio; CI, confidence interval.

Based on the KIR gene content and the Allele Frequency Net Database^24,25^, the KIR genetic profiles of the 55 MSM individuals were classified as either AA haplotypes, which carry only genes from the A haplotype (23.6%), or Bx haplotypes, which include both A and B haplotypes (AB) or only the B haplotype (BB) (76.4%)^31,32^.The high-risk group exhibited a significantly lower frequency of Bx haplotypes (50.00% vs. 85.37%, p = 0.007; OR = 5.833, 95% CI 1.498-22.711). In addition, a higher but not statistically significant frequency of AA haplotypes was detected in the same group (50.00% vs. 14.63%, p = 0.007; OR = 0.171 95% CI 0.044-0.667).

Haplotype diversity was analyzed using imputation from the *Allele Frequency Net Database*. Thirty-seven different multi-loci haplotypes were identified, of which two were AA and thirty-five were Bx. When comparing haplotype frequencies between the two MSM groups, only three haplotypes were shared (one AA and two Bx; see Supplementary Data 2). Notably, one AA haplotype (No.1 in Supplementary Data 2) was the most frequent overall (21.8%) and showed the largest difference between groups, being more frequent in the high-risk group (50% vs. 12%, p=0.007; OR=0.115, 95% CI 0.24-0.551). This haplotype is characterized by the absence of the 2DS2, 2DL2, 2DL5, 3DS1, 2DS5 and 2DS1 genes (see supplementary Data 2).

A linkage disequilibrium (LD) analysis was performed after excluding monomorphic genes (3DL3, 3DP1, 2DL4 and 3DL2). A higher frequency of LD among KIR genes was observed in the low-risk MSM group, although this difference may be influenced by the small sample size of this study, particularly in the high-risk group. The high-risk group showed strong LD between 2DS2-2DL2-2DS3 genes, not observed in the low-risk group. The significant p value linkage disequilibrium between genes is shown in Table 3.

**Table 3.**
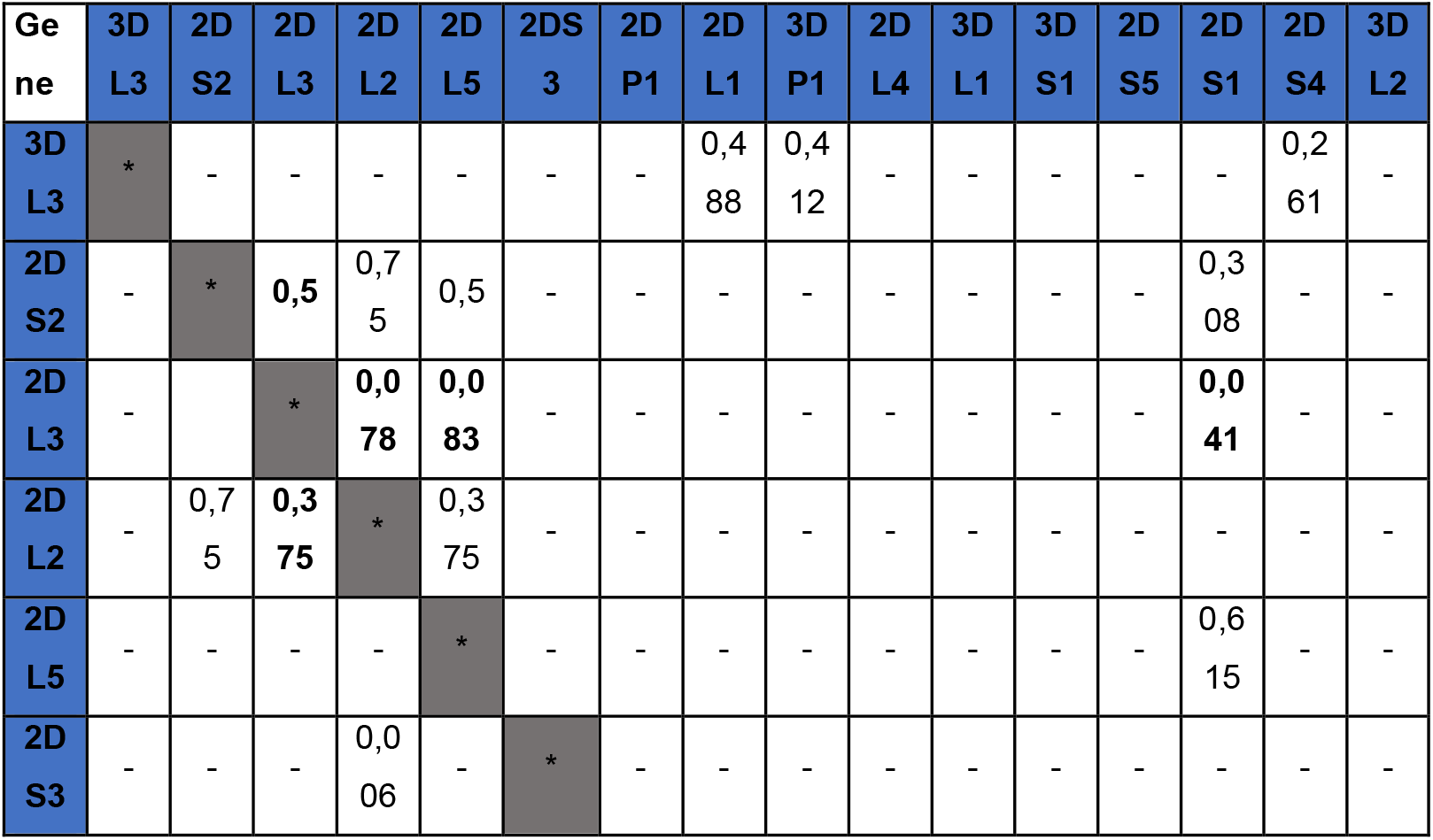
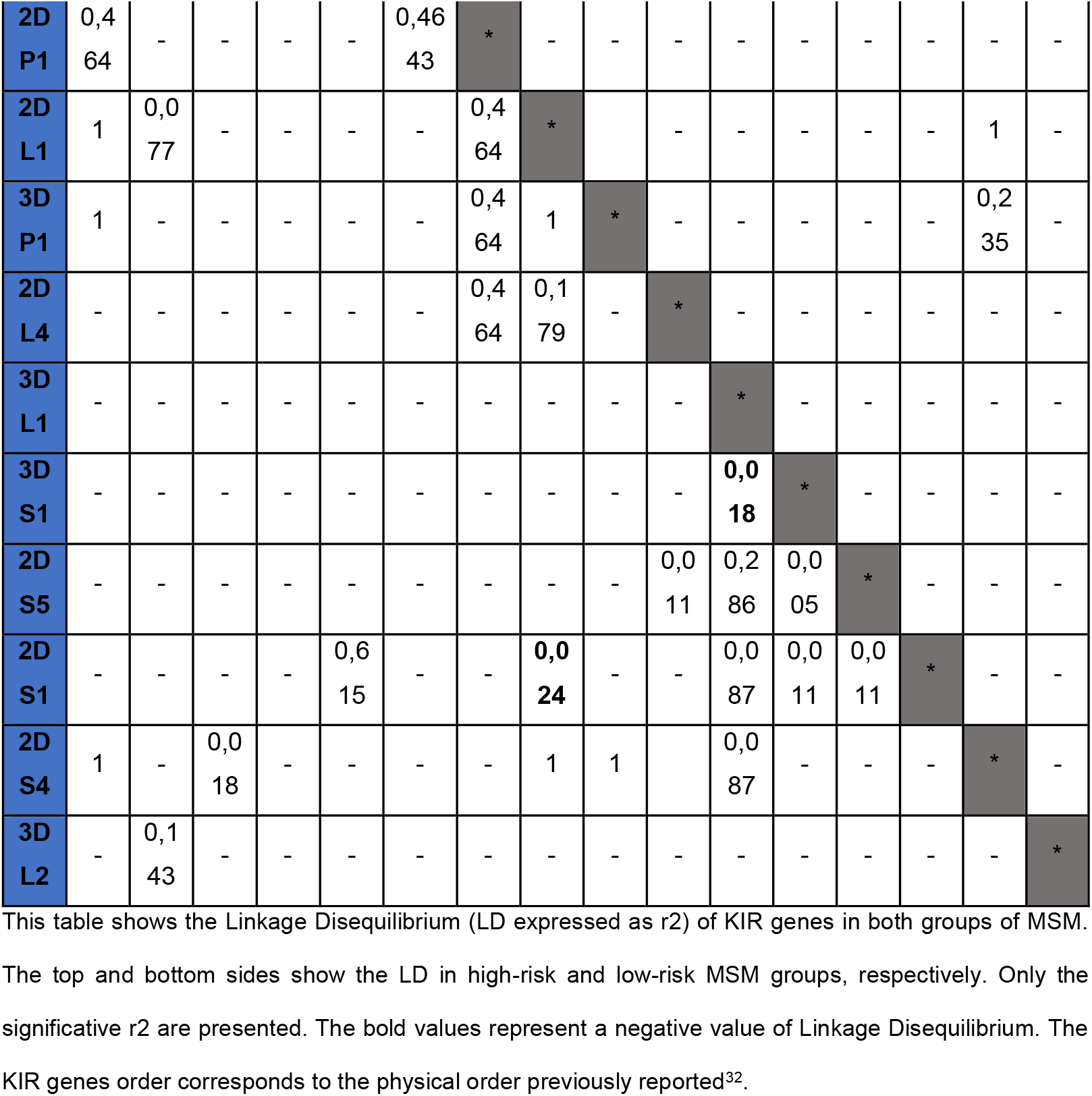
Linkage Disequilibrium of KIR genes in MSM groups.

Finally, we analyzed the previously reported association of the KIR3DL1/S1 and HLA-Bw4 combination with natural protection against HIV-1 infection. No significant differences were detected between the two MSM groups for these combinations (Table 2).

## Discussion

Here we demonstrated that compared with the low-risk group, the MSM in the high-risk group showed a protective genetic profile against the HIV-1 infection. This protective profile consisted of a higher frequency of the HLA-B*18 allele, a lower frequency of the HLA*B35, and an increased profile of inhibitory KIR genes, characterized for a low frequency of KIR2DL5, KIR2DS1, KIR2DS2 and KIR2DS5 genes and Bx haplotypes, and a higher frequency of a specific AA haplotype. Notably, the high-risk MSM group remains seronegative despite their risky sexual behaviors, which far exceed those associated with seroconversion in other MSM cohorts^29,33–35^. This situation suggests that these individuals have protective factors against HIV-1 infection that are independent of the CCR5 delta 32 mutation, as none of them carried this variant.

We found a significantly higher frequency of the HLA-B*18 allele in the high-risk MSM group. Several HLA alleles, most belonging to HLA-B, have been found at higher frequency in HESN compared to the general population and people living with HIV (PLWHIV), suggesting an association with the natural resistance to HIV-1^10^. The potential role of HLA-B*18 in HIV-1 acquisition is likely related to its function as a class I molecule involved in antigen presentation to CD8^+^ T cells, thereby contributing to early immune pressure on infected cells and viral control^36^. Farquhar et al. reported that HLA-B*18 allele was associated with a lower rate of HIV-1 mother-to-child transmission in Kenyan HIV+ mothers; likewise, no one of the HLA-B*18+ newborns were infected through breast milk^37^. This allele was also associated with HIV-1 natural resistance in serodiscordant couples from India, more frequent in HESN than their HIV-1 positive partners and healthy controls^38^. However, these findings have not been consistently replicated across populations or in recent studies, suggesting that its potential protective role may be context- and population-dependent.

We also showed a lower HLA-B*35 frequency in the high-risk MSM group, suggesting that this allele could be associated with HIV-1 protection. This result is consistent with previous studies showing that HLA-B*35 is associated with increased susceptibility to HIV-1 infection^39,40^. A study in India reported higher frequencies of HLA-B*35 in HIV+ positive individuals compared to healthy controls^41^. This allele has been associated with a significant risk of mother-to-child HIV-1 transmission. Arnaiz-Villena et al. demonstrated a higher frequency of this allele in HIV-1-infected children than in HIV-1-exposed but uninfected kids^42^. Furthermore, there is evidence that HLA-B*35+ individuals have a rapid disease progression to AIDS^40,42,43^. In a multicentric study, Carrington et al. observed an association of HLA-B*35 allele with rapid progression to AIDS in several cohorts of HIV+ infected individuals in the USA^44^. Recent structural and functional studies indicate that the impact of HLA-B35 on HIV-1 infection is driven by qualitative differences in peptide presentation and CD8^+^ T-cell responses. In particular, B35-Px variants exhibit reduced peptide-binding stability and impaired T-cell activation, which may contribute to suboptimal immune control of viral replication^45,46^.

Although the Bw4 allele and the heterozygosity of HLA have been associated with HIV-1 protection, we did not find differences in those or other previously described protective alleles between the MSM groups in our cohort^12,15,16^.

The protective effect against HIV-1 infection of some HLA molecules, including several HLA-B, has been related to a more remarkable ability to recognize essential HIV-1 antigens and induce a robust cytotoxic T lymphocyte (CTL) response against the virus^10,47,48^.

A lower frequency of KIR2DL5, KIR2DS1, and KIR2DS5 KIR genes was identified in the high-risk MSM group. This group exhibited a lower frequency of KIR-Bx haplotypes and a significantly increased frequency of one KIR-AA haplotype, which marked the most significant difference between the two groups of MSM. KIR2DL5, KIR2DS1, and KIR2DS5 genes are characteristic of the Bx haplotypes. They are absent in the AA haplotypes^32^, consistent with the low frequency of Bx haplotypes and the higher frequency of AA haplotypes observed in high-risk MSM. Importantly, this concordance between gene distribution and haplotype structure strongly supports the technical robustness and internal validity of the KIR genotyping performed in this study.

Various studies have suggested the protective role of some KIR genes during HIV-1 infection^16,49^. Protection has been commonly associated with a higher frequency of activating KIR genes, such as KIR3DS1, in HESN populations compared to PLHIV^16,24,50^. However, other studies have shown that inhibitory genes are also associated with protection^26^. Paximadis *et al*. reported that inhibitory genes KIR2DL2 and KIR2DL3 were more frequent in mother living with HIV who did not transmit the virus to their children compared to those transmitting mothers^51^. A meta-analysis that explored the influence of individual KIR genes and the genotypes KIR3DL1/KIR3DS1 in HIV acquisition in different populations demonstrated a robust protective effect against HIV-1 infection of both KIR2DL3 and KIR3DS1 polymorphisms in Caucasian, African and Asian HESN individuals^28^. In addition, a decreased risk of HIV-1 infection correlated with the presence of KIR2DL5, KIR2DS1 and KIR2DS5 genes in all studied populations^28^. These results differ from ours, where a lower frequency of those genes was associated with protection in the high-risk MSM group.

The KIR system is one of the most polymorphic regions of the human genome, and despite the considerable efforts toward their characterization, significant gaps remain in its identification and understanding^52^. Although extensive correlative studies show an association between some KIR alleles and haplotypes with HIV-1 infection, the broad variability between the methods to identify them, sample size, and genetic background of individuals enrolled in those studies contribute to the variability in the results regarding the KIR genes associated with susceptibility/protection to HIV-1 infection.

As previously described in the high-risk MSM group, the low frequency of KIR2DL5, KIR2DS1 and KIR2DS5 contrasts with the higher frequency of the AA haplotype. The AA haplotype encodes inhibitory KIRs with only one activating receptor (KIR2DS4)^52,53^. Although the Bx haplotype has been more frequently associated with low susceptibility to HIV-1^54^, a protective effect of the AA haplotype has been observed in other diseases, including classic Hodgkin lymphoma^47^ and during *Mycobacterium tuberculosis* infection^55^.

The balance of activating and inhibitory signals derived from the cell membrane receptors determines the NK cell responses. To become fully functional, NK cells must recognize self-HLA-I molecules through a self-specific KIR receptor in a process called cell licensing; thus, the NK cells that express inhibitory KIR are licensed, exhibiting a lower threshold for activation and more robust response upon further encounters with target cells^56,57^. Since the MSM in the high-risk group exhibited an inhibitory KIR profile, its protective effect against the HIV-1 infection may reside in the capacity of those receptors to generate a better licensing of NK cells^56,57^. In fact, the enrichment of KIR-AA haplotypes, which are predominantly composed of inhibitory KIR genes, may favor NK cell licensing through increased KIR–HLA interactions. Licensing mediated by inhibitory KIRs has been shown to enhance NK cell functional competence, including cytokine production and cytotoxicity^58,59^. In this context, inhibitory KIR engagement during NK cell development promotes functional responsiveness to target cells lacking self-HLA, a mechanism known as “missing-self” recognition, suggesting that this genetic profile could contribute to more effective antiviral responses^60^. Indeed, we previously observed a higher effector capacity of NK cells by IFN-γ production and increased cytotoxicity against K562 stimuli by MSM in the high-risk group^61,62^.

Additionally, this inhibitory genetic profile may also contribute to reduced basal immune activation, a feature consistently observed in HESN individuals and associated with decreased susceptibility to infection. Lower levels of immune activation may limit the availability of activated CD4+ T cells, the primary targets of HIV-1, thereby reducing the likelihood of productive infection despite repeated exposure^6,63^.

This study is highly informative since it describes genetic information in a population that is hard to enroll and follow, possibly exhibiting resistance to HIV-1 infection. Moreover, to our knowledge, this is the first report of KIR genes and multi-locus haplotypes in a Colombian population. However, several limitations should be considered. The small sample size, non-probabilistic sampling, and cross-sectional design limit the generalizability of our findings and preclude causal inferences. Given the exploratory nature of this study and the exceptional difficulty of recruiting HESN individuals with extreme risk behavior, we opted not to apply a stringent multiple-comparison correction (e.g., Bonferroni) to reduce the risk of type II errors; thus, the reported nominal p-values should be interpreted with caution.

Additionally, HLA typing was performed at moderate resolution, preventing the differentiation of alleles at the protein level, and KIR genotyping did not allow discrimination between KIR2DL5A and KIR2DL5B. This represents an important limitation, as moderate-resolution HLA typing does not enable the identification of specific allelic subtypes with potentially distinct functional properties. For example, different subtypes of HLA-B35 have been shown to exert divergent effects on HIV-1 disease progression, with B35-Px variants (such as B35:02 and B35:03) consistently associated with accelerated progression to AIDS, whereas B35-PY variants (such as B35:01 and B*35:08) appear to have minimal or no detrimental effect. Notably, these subtypes may differ by as little as a single amino acid yet result in markedly different immunological and clinical outcomes. Therefore, the associations reported in this study should be interpreted at the allele-group level, underscoring the importance of high-resolution HLA typing to refine immunogenetic analysis in HIV research^64^. We also did not analyze HLA-C alleles, nor assess HLA or KIR expression levels, and ancestry analysis was not performed to control for population stratification. These considerations suggest that our findings are hypothesis-generating and warrant validation in larger, independent cohorts with higher-resolution genotyping and functional studies.

## Conclusions

This study identifies HLA and KIR genetic patterns in Colombian MSM with high behavioral risk of HIV exposure who remain seronegative. The enrichment of HLA-B18, the reduced frequency of HLA-B35, and the predominance of KIR AA haplotypes suggest genetic signatures potentially associated with resistance to HIV infection in this population. These findings provide novel insights into the immunogenetic landscape of HIV susceptibility in Latin American populations and support further studies integrating larger cohorts and functional immune analyses to clarify the role of these genetic factors in natural resistance to HIV acquisition.

## Data Availability

All data produced in the present work are contained in the manuscript.

## Acknowledgments

The authors thank the volunteers for their kind participation, confidence, and priceless help. We also thank the Corporación Stonewall, Consejo Consultivo LGBTI de Medellín, Alianza Social LGBTI de Medellín y Antioquia, Fundación ANCLA, Fundación Antioqueña de Infectología (FAI), Red de Apoyo Social de Antioquia (RASA), and the Global Fund to Fight AIDS-Enterritorio, all institution from Medellín, Colombia, for their support in the consecution of volunteers and their training about working with vulnerable populations such as LGBTIQ+.

