## Supplementary material for "Potential association of HLA and KIR genetic profiles with resistance to HIV infection in high-risk men who have sex with men": Suplemental data 1 and 2

**Supplementary Data 1**

**HLA**‑**A and HLA**‑**B genotypes of HIV**‑**exposed seronegative MSM (n=60) stratified by risk group (high**‑**risk, n=16; low**‑**risk, n=44).**

| **Subject Code** | **Risk Group** | **HLA-A**   **Allele 1** | **HLA-A**  **Allele 2** | **HLA-B**  **Allele 1** | **HLA-B**  **Allele 2** |
| --- | --- | --- | --- | --- | --- |
| 002 | High-risk | 2 | 68 | 49 | 53 |
| 003 | High-risk | 23 | 34 | 65(14) | 44 |
| 004 | High-risk | 2 | 11 | 38 | 55 |
| 005 | Low-risk | 11 | 24 | 35 | 35 |
| 006 | High-risk | 24 | 68 | 61 | 53 |
| 007 | Low-risk | 24 | 24 | 61 | 48 |
| 008 | Low-risk | 24 | 24 | 62(15) | 35 |
| 009 | Low-risk | 1 | 1 | 8 | 58 |
| 010 | Low-risk | 2 | 24 | 39 | 61(40) |
| 011 | High-risk | 1 | 30 | 7 | 18 |
| 012 | Low-risk | 2 | 30 | 27 | 61(40) |
| 013 | Low-risk | 2 | 32 | 18 | 35 |
| 014 | Low-risk | 2 | 68 | 71(15) | 60(40) |
| 015 | Low-risk | 24 | 24 | 7 | 55 |
| 016 | Low-risk | 1 | 24 | 8 | 8 |
| 017 | High-risk | 29 | 30 | 65(14) | 60(40) |
| 018 | Low-risk | 24 | 26 | 35 | 35 |
| 019 | Low-risk | 2 | 3 | 61 | 50 |
| 020 | Low-risk | 1 | 33 | 14 | 57 |
| 021 | Low-risk | 11 | 24 | 35 | 52 |
| 022 | Low-risk | 26 | 29 | 38 | 45 |
| 023 | Low-risk | 3 | 33 | 35 | 57 |
| 024 | Low-risk | 2 | 23 | 44 | 44 |
| 025 | High-risk | 2 | 3 | 7 | 39 |
| 026 | Low-risk | 3 | 3 | 18 | 35 |
| 027 | High-risk | 3 | 11 | 64(14) | 39 |
| 028 | Low-risk | 3 | 24 | 61 | 51 |
| 029 | High-risk | 2 | 24 | 44 | 7 |
| 030 | Low-risk | 3 | 33 | 7 | 65(14) |
| 031 | Low-risk | 24 | 68 | 35 | 44 |
| 032 | Low-risk | 2 | 24 | 48 | 58 |
| 033 | Low-risk | 2 | 32 | 51 | 51 |
| 034 | Low-risk | 26 | 26 | 8 | 58 |
| 035 | Low-risk | 24 | 68 | 65(14) | 61(40) |
| 036 | Low-risk | 2 | _ | 35 | 61(40) |
| 037 | Low-risk | 23 | 30 | 8 | 44 |
| 038 | Low-risk | 26 | 33 | 38 | 41 |
| 039 | Low-risk | 2 | 3 | 44 | 51 |
| 040 | Low-risk | 24 | 24 | 61 | 35 |
| 041 | High-risk | 30 | 68 | 13 | 18 |
| 042 | High-risk | 2 | 24 | 62(15) | 44 |
| 043 | High-risk | 2 | 3 | 18 | 52 |
| 044 | High-risk | 2 | 24 | 18 | 35 |
| 045 | High-risk | 23 | 23 | 44 | 58 |
| 046 | Low-risk | 3 | 30 | 7 | 8 |
| 047 | Low-risk | 2 | 24 | 35 | 61(40) |
| 048 | Low-risk | 2 | 24 | 35 | 39 |
| 049 | Low-risk | 2 | 68 | 39 | 39 |
| 050 | Low-risk | 11 | 24 | 35 | 51 |
| 051 | High-risk | 2 | 3 | 53 | 58 |
| 052 | High-risk | 2 | 24 | 39 | 44 |
| 053 | Low-risk | 23 | 23 | 64(14) | 64(14) |
| 054 | Low-risk | 3 | 30 | 27 | 35 |
| 055 | Low-risk | 2 | 24 | 35 | 35 |
| 056 | Low-risk | 2 | 24 | 62(15) | 44 |
| 057 | Low-risk | 2 | 68 | 61(40) | 61(40) |
| 058 | Low-risk | 2 | 68 | 72(15) | 72(15) |
| 059 | Low-risk | 24 | 33 | 65(14) | 61(40) |
| 060 | Low-risk | 1 | 24 | 63(15) | 35 |
| 061 | Low-risk | 24 | 68 | 38 | 57 |

**Supplementary Data 1.** HLA‑A and HLA‑B genotypes of HIV‑exposed seronegative MSM (n=60) stratified by risk group (high‑risk, n=16; low‑risk, n=44).

For each subject, the two HLA‑A alleles and the two HLA‑B alleles are shown. Alleles are reported using standard nomenclature; numbers in parentheses indicate specific subtypes (e.g., 65(14) denotes HLA‑B*65:14). Missing data are indicated by an underscore (e.g., subject 036, HLA‑A allele 2). Genotypes were determined as described in the Methods section.

**Supplementary Data 2**

**KIR multi loci haplotypes and their frequency in high-risk and low-risk MSM**

| **Hap**. | **3DL3** | **2DS2** | **2DL3** | **2DL2** | **2DL5** | **2DS3** | **2DP1** | **2DL1** | **3DP1** | **2DL4** | **3DL1** | **3DS1** | **2DS5** | **2DS1** | **2DS4** | **3DL2** | **Group** | **All** | | **High-risk** | | **Low-risk** | |
| --- | --- | --- | --- | --- | --- | --- | --- | --- | --- | --- | --- | --- | --- | --- | --- | --- | --- | --- | --- | --- | --- | --- | --- |
|  |  |  |  |  |  |  |  |  |  |  |  |  |  |  |  |  |  | **Freq.** | **s.d.** | **Freq.** | **s.d.** | **Freq.** | **s.d.** |
| 1 | 1 | 0 | 1 | 0 | 0 | 0 | 1 | 1 | 1 | 1 | 1 | 0 | 0 | 0 | 1 | 1 | AA | 0.218 | 0.056 | 0.500 | 0.139 | 0.122 | 0.052 |
| 2 | 1 | 1 | 1 | 1 | 0 | 0 | 1 | 1 | 1 | 1 | 1 | 0 | 0 | 0 | 1 | 1 | Bx | 0.073 | 0.035 | 0.071 | 0.071 | 0.073 | 0.041 |
| 3 | 1 | 1 | 0 | 1 | 1 | 1 | 1 | 1 | 1 | 1 | 1 | 0 | 0 | 0 | 1 | 1 | Bx | 0.036 | 0.025 | 0.071 | 0.071 | 0.024 | 0.024 |
| 4 | 1 | 0 | 1 | ? | ? | 0 | 1 | 1 | 1 | 1 | 1 | 1 | 1 | ? | 1 | 1 | Bx | 0.018 | 0.018 | 0.071 | 0.071 |  |  |
| 5 | 1 | ? | 1 | 1 | 0 | ? | 1 | 1 | 1 | 1 | 1 | 0 | ? | 0 | ? | 1 | Bx | 0.018 | 0.018 | 0.071 | 0.071 |  |  |
| 6 | 1 | ? | 1 | ? | ? | ? | ? | ? | 1 | 1 | ? | 1 | ? | 0 | ? | 1 | Bx | 0.018 | 0.018 | 0.071 | 0.071 |  |  |
| 7 | 1 | 1 | 1 | 1 | 0 | 0 | 1 | 1 | 1 | 1 | ? | 0 | 0 | 0 | 1 | 1 | Bx | 0.018 | 0.018 | 0.071 | 0.071 |  |  |
| 8 | 1 | 1 | 0 | 1 | ? | 1 | 1 | 1 | 1 | 1 | 1 | 1 | 0 | 1 | 1 | 1 | Bx | 0.018 | 0.018 | 0.071 | 0.071 |  |  |
| 9 | 1 | 1 | 1 | 1 | 1 | 0 | 1 | 1 | 1 | 1 | 1 | 1 | 1 | 1 | 1 | 1 | Bx | 0.036 | 0.025 |  |  | 0.049 | 0.034 |
| 10 | 1 | 0 | 1 | 0 | 1 | 0 | 1 | 1 | 1 | 1 | 1 | 1 | 1 | 1 | 1 | 1 | Bx | 0.055 | 0.031 |  |  | 0.073 | 0.041 |
| 11 | 1 | 1 | 1 | 1 | ? | ? | 1 | 1 | 1 | 1 | ? | ? | 1 | ? | 1 | 1 | Bx | 0.018 | 0.018 |  |  | 0.024 | 0.024 |
| 12 | 1 | 1 | 1 | 1 | 1 | 0 | 1 | 1 | 1 | 1 | 1 | 0 | 1 | 1 | 1 | 1 | Bx | 0.018 | 0.018 |  |  | 0.024 | 0.024 |
| 13 | 1 | 1 | 1 | 1 | ? | 1 | 1 | 1 | 1 | 1 | 1 | 0 | 0 | 0 | 1 | 1 | Bx | 0.018 | 0.018 |  |  | 0.024 | 0.024 |
| 14 | 1 | 1 | 1 | 1 | 1 | 0 | 1 | 1 | 1 | 1 | 0 | 1 | 1 | 1 | ? | 1 | Bx | 0.018 | 0.018 |  |  | 0.024 | 0.024 |
| 15 | 1 | 1 | 1 | 1 | ? | 0 | 1 | 1 | 1 | 1 | 1 | 0 | 0 | 0 | 1 | 1 | Bx | 0.018 | 0.018 |  |  | 0.024 | 0.024 |
| 16 | 1 | 1 | 0 | 1 | 1 | ? | 1 | ? | 1 | 1 | ? | ? | 1 | 0 | ? | 1 | Bx | 0.018 | 0.018 |  |  | 0.024 | 0.024 |
| 17 | 1 | 1 | 1 | 1 | ? | ? | 0 | 1 | 1 | 1 | 1 | 0 | 0 | ? | 1 | 1 | Bx | 0.018 | 0.018 |  |  | 0.024 | 0.024 |
| 18 | 1 | 1 | 0 | 1 | ? | 0 | 0 | 0 | 1 | 1 | 1 | 1 | 1 | 1 | 1 | 1 | Bx | 0.018 | 0.018 |  |  | 0.024 | 0.024 |
| 19 | 1 | 1 | 0 | 1 | 0 | 0 | 1 | 1 | 1 | 1 | 1 | 0 | 0 | 0 | 1 | 1 | Bx | 0.018 | 0.018 |  |  | 0.024 | 0.024 |
| 20 | 1 | ? | 1 | ? | 1 | ? | 1 | ? | 1 | 1 | 1 | 1 | ? | ? | ? | 1 | Bx | 0.018 | 0.018 |  |  | 0.024 | 0.024 |
| 21 | 1 | 0 | 0 | 0 | 0 | 0 | 1 | 1 | 1 | 1 | 1 | 0 | 0 | 0 | 1 | 1 | AA | 0.018 | 0.018 |  |  | 0.024 | 0.024 |
| 22 | 1 | ? | 0 | ? | ? | 0 | 1 | ? | 1 | 1 | 0 | 1 | 1 | 1 | 0 | 1 | Bx | 0.018 | 0.018 |  |  | 0.024 | 0.024 |
| 23 | 1 | ? | ? | 1 | 1 | ? | 1 | 1 | 1 | 1 | 1 | 1 | ? | 1 | ? | 1 | Bx | 0.018 | 0.018 |  |  | 0.024 | 0.024 |
| 24 | 1 | 0 | 0 | 0 | ? | ? | 1 | 1 | 1 | 1 | 1 | 1 | 0 | ? | ? | 1 | Bx | 0.018 | 0.018 |  |  | 0.024 | 0.024 |
| 25 | 1 | 1 | 0 | 1 | 1 | 1 | 1 | 1 | 1 | 1 | 0 | 1 | 0 | 1 | ? | 1 | Bx | 0.018 | 0.018 |  |  | 0.024 | 0.024 |
| 26 | 1 | 1 | 1 | 1 | ? | ? | 1 | 1 | 1 | 1 | 1 | 0 | 0 | ? | 1 | 1 | Bx | 0.018 | 0.018 |  |  | 0.024 | 0.024 |
| 27 | 1 | ? | 1 | ? | ? | ? | 1 | ? | 1 | 1 | 1 | 1 | ? | 0 | 1 | 1 | Bx | 0.018 | 0.018 |  |  | 0.024 | 0.024 |
| 28 | 1 | 1 | 1 | 1 | ? | 1 | 1 | ? | 1 | 1 | 1 | 1 | 0 | 1 | 1 | 1 | Bx | 0.018 | 0.018 |  |  | 0.024 | 0.024 |
| 29 | 1 | 0 | 1 | 0 | 1 | 1 | 1 | 1 | 1 | 1 | 0 | 1 | 1 | 1 | 0 | 1 | Bx | 0.018 | 0.018 |  |  | 0.024 | 0.024 |
| 30 | 1 | 1 | 1 | 1 | 1 | 1 | 1 | 1 | 1 | 1 | 1 | 1 | 1 | 1 | 1 | 1 | Bx | 0.018 | 0.018 |  |  | 0.024 | 0.024 |
| 31 | 1 | 1 | 1 | 1 | 1 | 1 | 1 | 1 | 1 | 1 | 0 | 1 | 1 | 1 | 0 | 1 | Bx | 0.018 | 0.018 |  |  | 0.024 | 0.024 |
| 32 | 1 | 1 | ? | 1 | ? | 0 | ? | ? | 1 | 1 | ? | ? | 1 | 1 | ? | 1 | Bx | 0.018 | 0.018 |  |  | 0.024 | 0.024 |
| 33 | 1 | 1 | 1 | ? | 0 | 0 | 1 | 1 | 1 | 1 | 1 | 0 | 0 | 0 | 1 | 1 | Bx | 0.018 | 0.018 |  |  | 0.024 | 0.024 |
| 34 | 1 | 1 | 1 | ? | 1 | 0 | 1 | 1 | 1 | 1 | 1 | 1 | 1 | 1 | 1 | 1 | Bx | 0.018 | 0.018 |  |  | 0.024 | 0.024 |
| 35 | 1 | 1 | 0 | 1 | 1 | 1 | 1 | 1 | 1 | 1 | 1 | 1 | 0 | 1 | 1 | 1 | Bx | 0.018 | 0.018 |  |  | 0.024 | 0.024 |
| 36 | 1 | 1 | 0 | 1 | 1 | 0 | 0 | 0 | 1 | 1 | 1 | 1 | 1 | 1 | 1 | 1 | Bx | 0.018 | 0.018 |  |  | 0.024 | 0.024 |
| 37 | 1 | 1 | 1 | ? | 1 | 0 | 1 | 1 | 1 | 1 | 1 | 0 | 1 | 1 | 1 | 1 | Bx | 0.018 | 0.018 |  |  | 0.024 | 0.024 |

Supplementary Data 2. KIR multi‑locus haplotypes and their frequencies in high‑risk (n=14) and low‑risk (n=41) HIV‑exposed seronegative MSM.

For each of the 37 haplotypes identified, the presence (1) or absence (0) of the indicated KIR genes is shown. Question marks (?) denote missing data. Haplotypes are classified as AA (carrying only A haplotype genes) or Bx (including A and/or B haplotype genes). For each haplotype, the overall frequency (Freq.), standard deviation (s.d.), and frequencies within the high‑risk and low‑risk groups are provided. The total number of individuals analyzed was 55. Haplotype frequencies may not sum to 100% due to rounding and missing data. See Methods for haplotype imputation details.
